# Change in cognition and body mass index in relation to preclinical dementia

**DOI:** 10.1101/2021.02.01.21250918

**Authors:** Ida K. Karlsson, Yiqiang Zhan, Margaret Gatz, Chandra A. Reynolds, Anna K. Dahl Aslan

## Abstract

**INTRODUCTION:** To study if declining cognition drives weight loss in preclinical dementia, we examined the longitudinal association between body mass index (BMI) and cognitive abilities in those who did or did not later develop dementia.

**METHODS:** Using data from individuals spanning age 50-89, we applied dual change score models separately in individuals who remained cognitively intact (n=1,498) and those who were diagnosed with dementia within five years of last assessment (n=459).

**RESULTS:** Among the cognitively intact, there was a bidirectional association: stable BMI predicted stable cognition and vice versa. Among those subsequently diagnosed with dementia, the association was unidirectional: higher BMI predicted declining cognition, but cognition did not predict change in BMI.

**DISCUSSION:** While BMI and cognition stabilized each other when cognitive functioning was intact, this buffering effect was missing in the preclinical dementia phase. This finding indicates that weight loss in preclinical dementia is not driven by declining cognition.

## 1. Background

Pathophysiological changes in dementia start many years before clinical manifestations are seen. During this preclinical phase, cognitive abilities decline progressively, and are followed by a decline in functional abilities[1]. Unintentional weight loss appears common during the preclinical dementia phase[7]. This weight loss is hypothesized to explain the “obesity paradox”, where a high body mass index (BMI) in midlife has been robustly and positively associated with cognitive decline and dementia, while a higher BMI in late-life may instead be associated with lower dementia risk[3-5].

BMI generally increases from early adulthood through age 65, after which it levels off to then start to decrease around age 80[6]. These fluctuations appear to be more pronounced among individuals who are later diagnosed with dementia, where BMI tends to increase more at earlier ages, but then starts to fall more sharply approximately 10 years prior to diagnosis[7]. It is thus plausible that BMI can act both as a risk factor and a prodromal sign of dementia, depending on the timing of measurement and on longitudinal weight trajectories.

Importantly, the associations between BMI and cognitive abilities may differ in normative cognitive aging and preclinical dementia, where it is not established whether declining cognitive abilities are a driver of weight loss. To investigate this question, we aimed to study the longitudinal and dynamic association between BMI and cognitive abilities, including the direction of effect, separately in individuals who later developed dementia and those who remained cognitively intact. We thereby hope to better understand the complex relationship between overweight, cognitive abilities, and dementia, and the nature of the obesity paradox.

## 2. Methods

### 2.1 Study population

We used data from three longitudinal studies of aging within the Swedish Twin Registry[8], with rich longitudinal cognitive data, dementia diagnoses, and linkages to healthcare registers, making them ideal for the questions under study. The Swedish Adoption/Twin Study of Aging (SATSA)[9] consists of 859 individuals from same-sex twin pairs, who participated in up to 10 in-person testing phases between 1986 and 2014. Aging in Women and Men (GENDER)[10] includes 496 individuals from 248 opposite-sex twin pairs, who participated in up to three in-person testing phases conducted on a four-year rolling schedule between 1995 and 2005. Origins of Variance in the Oldest Old: Octogenarian Twins (OCTO-Twin)[11] consists of 702 individuals from 351 same-sex twin pairs over age 80, who participated in up to five in-person testing occasions conducted on a two-year rolling schedule between 1991 and 2001. The in-person testing phases were conducted in a similar manner across the three studies, and included a health examination, cognitive assessments, and an extensive interview. We could thus pool individuals from the three studies, yielding a study sample of 2,057 individuals.

All participants provided informed consent, and the studies were approved by the Regional Ethics Board in Stockholm.

### 2.2 BMI measurements

Height and weight were measured by trained research nurses as part of the health examinations during each in-person testing occasion. The measures have been thoroughly examined for outliers, both quantitatively and visually by plotting individual trajectories over time, as described in detail previously[12]. BMI was calculated as kilograms divided by height (in meters) squared. BMI measures below 15 or above 55 and unrealistic changes over a short time period were set to missing, but BMI was otherwise allowed to vary. Individual BMI trajectories across age showed comparable patterns in the three studies (Figure S1).

### 2.3 Cognitive measures

During each testing occasion, cognitive tests were performed covering four domains: verbal abilities (Synonyms), spatial abilities (Block Design), episodic memory (Thurstone’s Picture Memory Task), and processing speed (Symbol Digit)[13]. Principal component analysis, based on the individual tests, was done to create a measure of general cognitive ability, which was standardized relative to means and variances in the first testing occasion[14]. All cognitive measures were transformed into T-scores (mean 50 and standard deviation of 10) prior to analyses, scaled to the first in-person testing occasion. General cognitive ability, spatial ability, episodic memory, and processing speed were included in this study. Verbal ability could not be included due to issues with model convergence. Individual trajectories were comparable across studies for all cognitive measures (Figure S1).

### 2.4 Dementia status

All studies entailed a dementia evaluation, and, in addition, dementia after the end of study was retrieved from nationwide healthcare registers (the National Patient Register, Cause of Death Register, and Prescribed Drug Register), as described in detail previously[12]. Individuals diagnosed with dementia either during or within five years of last study participation were categorized into the preclinical dementia group; those not diagnosed during that period were categorized as cognitively intact. For analysis, we used only cognitive measures prior to dementia diagnosis.

### 2.5 Covariates

Information about age at each testing occasion, sex, and education was available. Education was categorized into seven years or less or more than seven years, corresponding to basic or more than basic education at the time.

### 2.6 Statistical analyses

Dual change score models (DCSMs)[15, 16] were conducted in Mplus[17] to study whether level of BMI predicts change in cognitive abilities, and whether level of cognitive abilities predict change in BMI.

The data were first split into 2-year age intervals according to age when BMI and cognitive ability were measured (age 50-51 through 88-89). Sex and education were adjusted for by regressing them on intercept levels and slopes, and age (in two-year bins) used as the underlying timescale. Relatedness between twins was accounted for with robust standard errors. All model comparisons described below were done by applying the log-likelihood difference test with an maximum likelihood robust (MLR) correction for scaling factors [18]. Statistical significance threshold was set at α=0.05.

A path-diagram describing the DCSM model along with formulas for calculating change is provided in Figure 1. Univariate DCSMs of linear and non-linear change in BMI and cognitive abilities were first applied. In addition to estimating the mean intercept level (μ_BMIi_, μ_COGi_) and linear slope (μ_BMIs,_ μ_COGs_), the model estimates change from one time point to the next (Δ_BMIt_ and Δ_COGt_) as a function of a static linear slope (BMI_S_, COG_S_) plus a proportional change (β_BMI_, β_COG_), which is relative to the previous level and thus a measure of non-linear change from one time point to the next.

**Figure 1:**
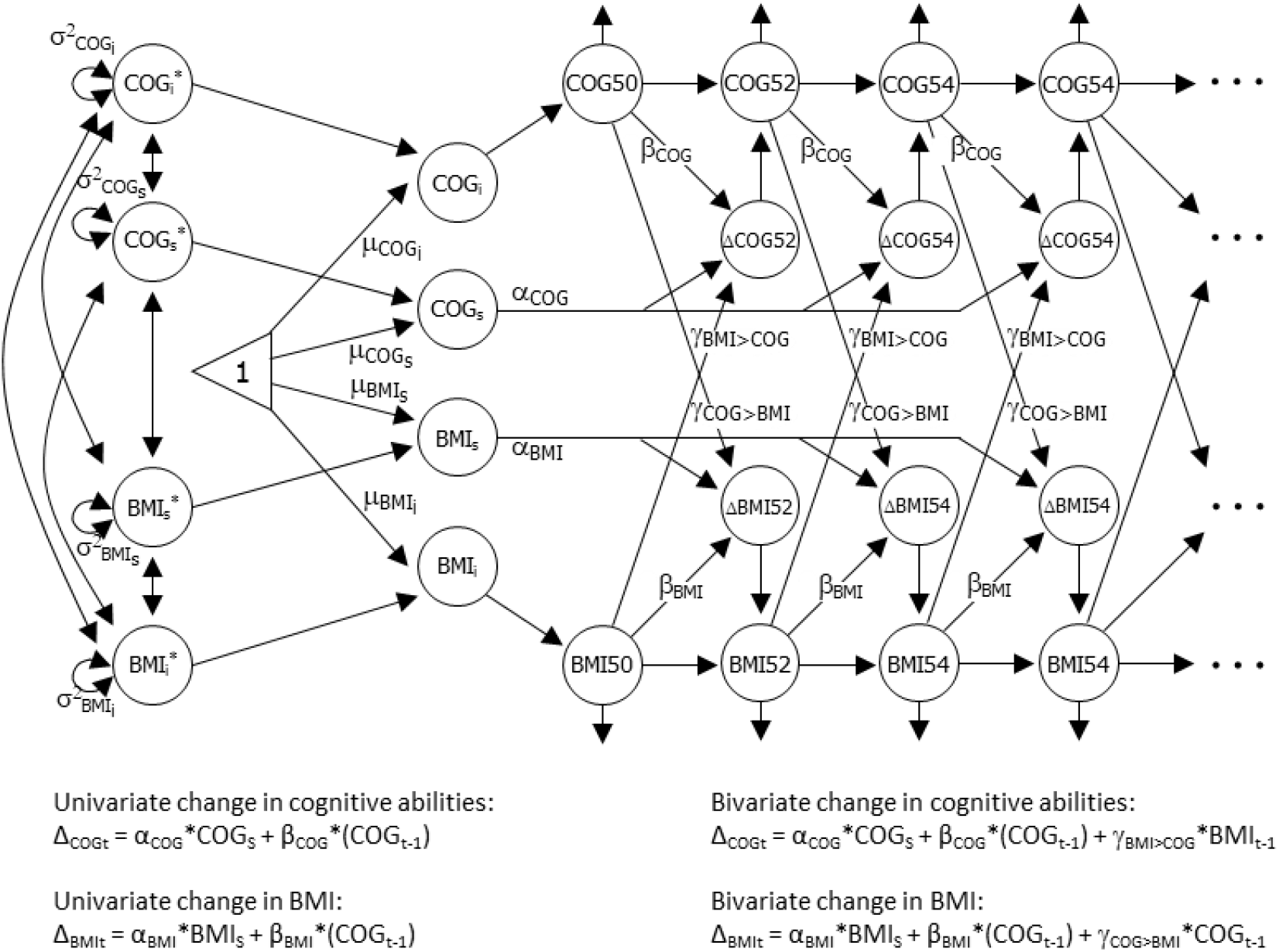
Path diagram of the bivariate dual change score model of BMI and cognition. Level of BMI and cognitive abilities (COG) are modeled in each age category (BMI50, BMI52…; COG50, COG52…). BMI_i_, BMI_S_, COG_i_, and COG_S_ represent intercept level and slope of BMI and cognitive ability, μ_BMI50_, μ_BMIS_, μ_COG50_, and μ_COGS_ their estimated mean levels, and σ^2^_BMIi_, σ^2^_BMIs_, σ^2^_COGi_, and σ^2^_COGs_ their variances. α_BMI_ and α_COG_ represent the constant change and is fixed at 1, while β_BMI_ and β_COG_ represent the proportional change from one time point to the next. The coupling effect of BMI on cognition is represented by the γ_BMI>COG_ parameter, and that of cognition on BMI by the γ_COG>BMI_ parameter. The equations on the left show the univariate change in cognition and BMI, respectively. Univariate change in BMI at age=_t,_ is here determined by the constant change (α_BMI_ * BMI_S_) plus the proportional change multiplied by BMI level at the preceding time point (β_BMI_ * BMI_t-1_). When a breakpoint is included in the model, the β_BMI_ parameter can differ before and after the breakpoint. The equations on the right show bivariate change in cognition and BMI, respectively. For change in BMI, the effect of cognitive ability is considered by adding the coupling effect which is multiplied by cognitive ability at the preceding time point to the formula (γ_COG>BMI_ * COG_t-1_). As with the β_BMI_ parameter, the γ_COG>BMI_ parameter can differ before and after a breakpoint. Univariate and bivariate change in cognitive abilities is determined by the corresponding formulas and parameters.

The linear slope and proportional change are assumed to be constant over time. To test for differences in rate of change, we included breakpoints in the univariate models after age 65, 69, and 75, which allows for different proportional change before and after the breakpoint. The best fitting breakpoint-model was selected based on Akaike information criteria (AIC), and compared to a null-model without a breakpoint.

Next, bivariate DCSMs were applied, modelling the dynamic association between BMI and cognitive abilities. In addition to the parameters in the univariate DCSM, the bivariate model also estimates cross-trait coupling parameters. These coupling parameters are estimates of how change in cognition from one time point to the next is influenced by BMI level at the previous time point (γ_BMI>COG_), and vice versa (γ_COG>BMI_). As with the univariate proportional parameters, these coupling parameters are assumed to be constant across time but can differ before and after a breakpoint.

By comparing models with and without the coupling parameters, the temporal order of changes can be tested. First, a full-coupling (bidirectional) model, with both coupling parameters included, was applied. This was compared to a no-coupling model with neither coupling parameter included to test for any association, and to two models with only one of the coupling parameters to test for unidirectional effects. These model comparisons were carried out both in the full sample and by dementia status.

Lastly, we tested for differences in specific model parameters by dementia status. We first examined group-differences in univariate parameters, by comparing a model where all univariate parameters were allowed to vary freely in the two groups to models where 1) residual variances, 2) variances and covariances, 3) proportional change parameter, and 4) mean intercept and slope were constrained in a stepwise manner. We then examined group-differences in bivariate parameters in a similar manner, by comparing a model where all univariate and bivariate parameters were free to vary freely across groups to models where 1) coupling parameters, 2) residual covariance, and 3) cross-trait covariance between intercepts and slopes (i.e. all bivariate parameters) were constrained in a stepwise manner.

## 3. Results

### 3.1 Study population

Among the 2,057 study participants, 1,959 had measures of BMI and/or cognition between age 50 and 89. Two individuals were excluded due to missing information about education, leaving a final analysis sample of 1,957 individuals. Stratifying on dementia status yielded 1,498 individuals in the cognitively intact group, and 459 individuals in the group diagnosed with dementia within five years of last cognitive measurement. Sample characteristics are presented in Table 1.

**Table 1:** Descriptive statistics of the full sample and stratified by dementia status.

|  | **Full sample** | **Cognitively intact** | **Dementia** |
| --- | --- | --- | --- |
| Individuals, N | 1,957 | 1,498 | 459 |
| Age at baseline, mean (range; SD) | 72.5 (50.1-89.9; 10.2) | 71.4 (50.1-89.9; 10.5) | 76.0 (51.6-89.8; 8.3) |
| Follow-up time, mean (range; SD) | 8.0 (0-27.0; 7.3) | 8.3 (0-27.0; 7.5) | 7.1 (0-26.2; 6.8) |
| Testing occasions, mean (range; SD) | 3.4 (1-10; 2.2) | 3.5 (1-10; 2.2) | 3.1 (0-9; 1.9) |
| Women, N (%) | 1,155 (59%) | 287 (62.5) | 868 (57.9) |
| Lower education, N (%) | 1,107 (57%) | 293 (63.8) | 814 (54.3) |

*Note.* N: number; SD: standard deviation
Descriptive statistics for the full sample, and separately for individuals who remain cognitively intact and those who are diagnosed with dementia during or within 5 years after the study. The number (%) of individuals are presented for categorical variables and the mean level (range and standard deviation) for continuous variables.

### 3.1 Univariate trajectories of BMI and cognitive abilities

Models allowing for non-linear age trajectories had a significantly better fit for all outcomes (p≤0.01). For BMI, a model with a breakpoint at age 69 had the best fit, and for all cognitive domains, a breakpoint at age 65 had the best fit. These breakpoints were selected for all further analyses of respective trait.

#### 3.1.1 Full sample

The full univariate model estimates are presented in Table S1. The predicted mean BMI level at age 50 was 25.19 in the full sample, with a negative overall linear slope (μ_BMIS_=-1.337). Significant buffering effects (β_BMI<69_=0.058, β_BMI>69_=0.045) indicate that higher BMI predicted steeper increase from age 50 to 69, and less decline from age 69 to 89 (Table S1, univariate; Figure 2 left panel, dashed lines).

**Figure 2:**
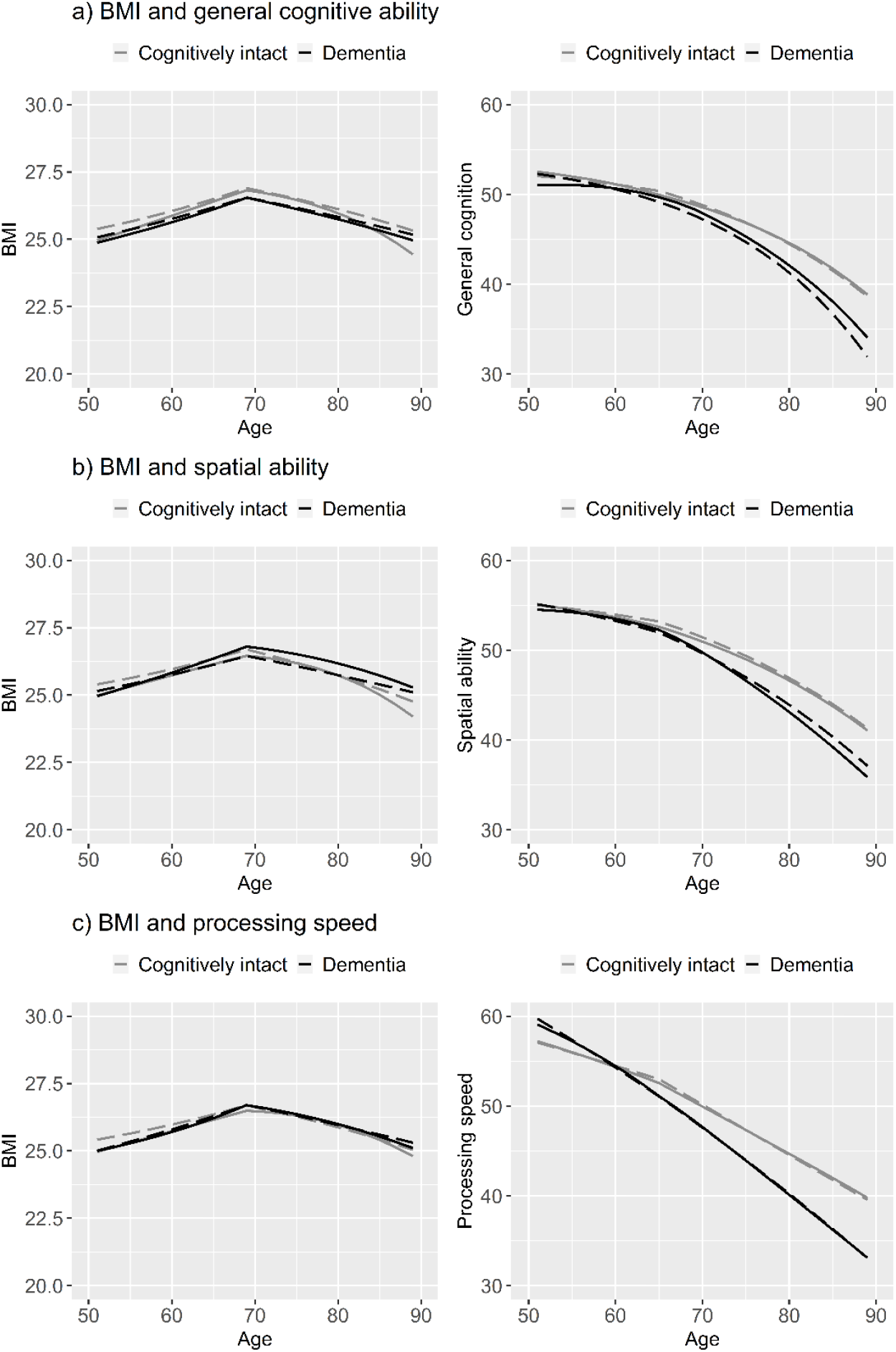
Longitudinal trajectories of change in body mass index and cognitive abilities from age 50-89, with and without the bivariate coupling-parameter. Trajectories from the full coupling dual change score model are shown in solid lines, and those from the no-coupling dual change score model in dashed lines. Trajectories of individuals who remain cognitively intact are shown in grey, and those who were diagnosed with dementia within 5 years after last cognitive measure in black. Models were adjusted for sex and education, and a breakpoint was included to allow for differences in the proportional and coupling effects before and after age 69 in the trajectories of BMI and after age 65 in the trajectories of cognitive ability.

Mean general cognitive ability at age 50 was 51.901 (Table S1a, univariate). There was a negative linear slope (μ_BMIS_=-5.739), but with buffering from proportional effects (β_COG<65_=0.107, β_COG>65_=0.103) resulting in a slight decline from age 50 to 65, followed by a steeper decline from age 65 to 89 (Figure 2a right panel, dashed lines). As proportional effects are multiplied with the cognitive level at the preceding occasion, it means that higher cognitive ability predicted less decline. It is also important to note that this means that declining cognitive ability can be accelerated due to increasingly weaker buffering effects with lower cognitive levels. Pattern of change in spatial ability was similar to that in general cognitive ability (Figure 2b right panel, dashed lines; Table S1b, univariate). Change in episodic memory was driven by a strongly negative linear slope, but with substantial buffering from the proportional effects leading to stable levels from age 50 through 65, followed by a gradual decline after 65 (Table S1c, univariate). Processing speed had little buffering from proportional effects and declined already between age 50 and 64, after which decline was more substantial (Figure 2d right panel, dashed lines; Table S1d, univariate).

#### 3.1.1 By dementia status

Compared to those who remained cognitively intact, individuals who were later diagnosed with dementia had significantly lower predicted BMI (estimate difference 0.244 at age 50; Table 2, univariate), but did not differ in terms of longitudinal rate of change in BMI (mean slope or proportional effects, Table 2, univariate).

**Table 2:** Univariate and bivariate change in body mass index and cognitive abilities from age 50-89, stratified by dementia status.

| a) <u>General cognitive ability</u> | <u>Cognitively intact</u> |  |  |  | <u>Dementia</u> |  |  |  | <u>Group difference, LRT</u> |  |  |
| --- | --- | --- | --- | --- | --- | --- | --- | --- | --- | --- | --- |
|  | <u>Univariate</u> |  | <u>Bivariate</u> |  | <u>Univariate</u> |  | <u>Bivariate</u> |  | -2LL | DGF | P-value |
|  | Est | SE | Est | SE | Est | SE | Est | SE |  |  |  |
| **BMI parameters** |  |  |  |  |  |  |  |  |  |  |  |
| Mean intercept level, age 50 ($\mu_{BMI_i}$) | 25.276 | 0.315* | 24.963 | 0.345* | 25.032 | 0.755* | 24.874 | 0.742* | 9.332 | 1 | 0.002<sup>a</sup> |
| Mean slope ($\mu_{BMI_s}$) | -1.339 | 0.391* | -2.465 | 0.600* | -0.610 | 0.935 | -0.33 | 0.821 | 0.003 | 1 | 0.956<sup>a</sup> |
| Proportional effect <69 ($\beta_{BMI <69}$) | 0.058 | 0.015* | 0.054 | 0.016* | 0.030 | 0.036 | 0.034 | 0.037 | 0.475 | 2 | 0.789<sup>a</sup> |
| Proportional effect >69 ($\beta_{BMI >69}$) | 0.045 | 0.015* | 0.030 | 0.015* | 0.018 | 0.036 | 0.000 | 0.034 | 0.475 | 2 | 0.789<sup>a</sup> |
| **Cognition parameters** |  |  |  |  |  |  |  |  |  |  |  |
| Mean intercept level ($\mu_{COG_i}$) | 52.037 | 0.697* | 52.518 | 0.688* | 52.555 | 1.771* | 51.045 | 1.909* | 159.788 | 1 | <0.001<sup>a</sup> |
| Mean slope ($\mu_{COG_s}$) | -5.139 | 1.006* | -6.432 | 1.502* | -6.631 | 1.736* | 3.480 | 6.193 | 2.891 | 1 | 0.089<sup>a</sup> |
| Proportional effect <65 ($\beta_{COG <65}$) | 0.096 | 0.020* | 0.125 | 0.021* | 0.120 | 0.036* | 0.124 | 0.042* | 0.092 | 2 | 0.955<sup>a</sup> |
| Proportional effect >65 ($\beta_{COG >65}$) | 0.091 | 0.021* | 0.096 | 0.020* | 0.120 | 0.039* | 0.144 | 0.035* | 0.092 | 2 | 0.955<sup>a</sup> |
| **Bivariate parameters** |  |  |  |  |  |  |  |  |  |  |  |
| Coupling cognition → BMI <69 ($\gamma_{COG \rightarrow BMI, <69}$) | - | - | 0.025 | 0.011* | - | - | -0.007 | 0.014 | 11.133 | 2 | 0.004<sup>b</sup> |
| Coupling cognition → BMI >69 ($\gamma_{COG \rightarrow BMI, >69}$) | - | - | 0.032 | 0.010* | - | - | 0.004 | 0.013 | 11.133 | 2 | 0.004<sup>b</sup> |
| Coupling BMI → cognition <65 ($\gamma_{BMI \rightarrow COG, <65}$) | - | - | -0.015 | 0.049 | - | - | -0.393 | 0.254 | 27.947 | 2 | <0.001<sup>b</sup> |
| Coupling BMI → cognition >65 ($\gamma_{BMI \rightarrow COG, >65}$) | - | - | 0.042 | 0.049 | - | - | -0.429 | 0.25 | 27.947 | 2 | <0.001<sup>b</sup> |

| b) <u>Spatial ability</u> | <u>Cognitively intact</u> |  |  |  | <u>Dementia</u> |  |  |  | <u>Group difference, LRT</u> |  |  |
| --- | --- | --- | --- | --- | --- | --- | --- | --- | --- | --- | --- |
|  | <u>Univariate</u> |  | <u>Bivariate</u> |  | <u>Univariate</u> |  | <u>Bivariate</u> |  | -2LL | DGF | P-value |
|  | Est | SE | Est | SE | Est | SE | Est | SE |  |  |  |
| **BMI parameters** |  |  |  |  |  |  |  |  |  |  |  |
| Mean intercept level, age 50 ($\mu_{BMI_i}$) | 25.276 | 0.315* | 24.986 | 0.339* | 25.032 | 0.755* | 24.961 | 0.751* | 9.332 | 1 | 0.002<sup>a</sup> |
| Mean slope ($\mu_{BMI_s}$) | -1.339 | 0.391* | -2.621 | 0.708* | -0.610 | 0.935 | -0.439 | 0.967 | 0.003 | 1 | 0.956<sup>a</sup> |
| Proportional effect <69 ($\beta_{BMI <69}$) | 0.058 | 0.015* | 0.052 | 0.018* | 0.030 | 0.036 | 0.025 | 0.037 | 0.475 | 2 | 0.789<sup>a</sup> |
| Proportional effect >69 ($\beta_{BMI >69}$) | 0.045 | 0.015* | 0.025 | 0.016 | 0.018 | 0.036 | -0.011 | 0.035 | 0.475 | 2 | 0.789<sup>a</sup> |
| **Cognition parameters** |  |  |  |  |  |  |  |  |  |  |  |
| Mean intercept level ($\mu_{COG_i}$) | 55.177 | 0.714* | 55.045 | 0.780* | 54.49 | 1.491* | 54.518 | 1.873* | 67.557 | 1 | <0.001<sup>a</sup> |
| Mean slope ($\mu_{COG_s}$) | -4.41 | 0.756* | -4.832 | 1.200* | -3.866 | 0.881* | 3.537 | 3.350 | 0.571 | 1 | 0.450<sup>a</sup> |
| Proportional effect <65 ($\beta_{COG <65}$) | 0.076 | 0.014* | 0.096 | 0.020* | 0.065 | 0.016* | 0.064 | 0.040 | 0.981 | 2 | 0.612<sup>a</sup> |
| Proportional effect >65 ($\beta_{COG >65}$) | 0.070 | 0.015* | 0.075 | 0.019* | 0.058 | 0.019* | 0.073 | 0.030* | 0.981 | 2 | 0.612<sup>a</sup> |
| **Bivariate parameters** |  |  |  |  |  |  |  |  |  |  |  |
| Coupling cognition → BMI <69 ($\gamma_{COG \rightarrow BMI, <69}$) | - | - | 0.027 | 0.015 | - | - | 0.000 | 0.018 | 3.73 | 2 | 0.155<sup>b</sup> |
| Coupling cognition → BMI >69 ($\gamma_{COG \rightarrow BMI, >69}$) | - | - | 0.037 | 0.015* | - | - | 0.013 | 0.017 | 3.73 | 2 | 0.155<sup>b</sup> |
| Coupling BMI → cognition <65 ($\gamma_{BMI \rightarrow COG, <65}$) | - | - | -0.028 | 0.046 | - | - | -0.286 | 0.144* | 12.581 | 2 | 0.002<sup>b</sup> |
| Coupling BMI → cognition >65 ($\gamma_{BMI \rightarrow COG, >65}$) | - | - | 0.01 | 0.042 | - | - | -0.313 | 0.129* | 12.581 | 2 | 0.002<sup>b</sup> |

| c) <u>Processing speed</u> | <u>Cognitively intact</u> |  |  |  | <u>Dementia</u> |  |  |  | <u>Group difference, LRT</u> |  |  |
| --- | --- | --- | --- | --- | --- | --- | --- | --- | --- | --- | --- |
|  | Univariate |  | Bivariate |  | Univariate |  | Bivariate |  | -2LL | DGF | P-value |
|  | Est | SE | Est | SE | Est | SE | Est | SE |  |  |  |
| **BMI parameters** |  |  |  |  |  |  |  |  |  |  |  |
| Mean intercept level, age 50 ($\mu_{BMIi}$) | 25.276 | 0.315* | 24.966 | 0.335* | 25.032 | 0.755* | 25.002 | 0.768* | 9.332 | 1 | 0.002<sup>a</sup> |
| Mean slope ($\mu_{BMIs}$) | -1.339 | 0.391* | -2.137 | 0.549* | -0.610 | 0.935 | -0.586 | 0.95 | 0.003 | 1 | 0.956<sup>a</sup> |
| Proportional effect <69 ($\beta_{BMI <69}$) | 0.058 | 0.015* | 0.058 | 0.016* | 0.030 | 0.036 | 0.043 | 0.034 | 0.475 | 2 | 0.789<sup>a</sup> |
| Proportional effect >69 ($\beta_{BMI >69}$) | 0.045 | 0.015* | 0.030 | 0.015 | 0.018 | 0.036 | 0.002 | 0.034 | 0.475 | 2 | 0.789<sup>a</sup> |
| **Cognition parameters** |  |  |  |  |  |  |  |  |  |  |  |
| Mean intercept level ($\mu_{COGi}$) | 56.882 | 0.850* | 57.216 | 0.862* | 59.869 | 2.325* | 59.076 | 2.250* | 47.991 | 1 | <0.001<sup>a</sup> |
| Mean slope ($\mu_{COGs}$) | -1.098 | 0.669* | -2.043 | 1.071 | -2.153 | 0.971* | 2.389 | 3.574 | 19.49 | 1 | <0.001<sup>a</sup> |
| Proportional effect <65 ($\beta_{COG <65}$) | 0.009 | 0.013 | 0.030 | 0.014 | 0.016 | 0.018 | 0.043 | 0.032 | 0.868 | 2 | 0.648<sup>a</sup> |
| Proportional effect >65 ($\beta_{COG >65}$) | 0.000 | 0.014 | 0.002 | 0.014 | 0.015 | 0.022 | 0.025 | 0.022 | 0.868 | 2 | 0.648<sup>a</sup> |
| **Bivariate parameters** |  |  |  |  |  |  |  |  |  |  |  |
| Coupling cognition → BMI <69 ($\gamma_{COG>BMI, <69}$) | - | - | 0.015 | 0.008 | - | - | -0.006 | 0.013 | 5.076 | 2 | 0.079<sup>b</sup> |
| Coupling cognition → BMI >69 ($\gamma_{COG>BMI, >69}$) | - | - | 0.026 | 0.008* | - | - | 0.009 | 0.012 | 5.076 | 2 | 0.079<sup>b</sup> |
| Coupling BMI → cognition <65 ($\gamma_{BMI>COG, <65}$) | - | - | -0.011 | 0.039 | - | - | -0.233 | 0.131 | 5.816 | 2 | 0.055<sup>b</sup> |
| Coupling BMI → cognition >65 ($\gamma_{BMI>COG, >65}$) | - | - | 0.034 | 0.038 | - | - | -0.19 | 0.146 | 5.816 | 2 | 0.055<sup>b</sup> |

Note. -2LL: -2 log-likelihood; BMI: body mass index; COG: cognition; DGF: degrees of freedom; Est: estimate; LRT: likelihood ratio test; SE: standard error
\* $p < 0.05$ ; <sup>a</sup> tested in univariate models; <sup>b</sup> tested in bivariate models
Parameter estimates, standard deviations, and p-values from the univariate and bivariate dual change score models of BMI and cognitive abilities, separately for individuals who remain cognitively intact and those who are diagnosed with dementia during or within 5 years after the study. Models are adjusted for sex and education. A breakpoint was included to allow for differences in the proportional change before and after age 69 in the trajectories of BMI and after age 65 in the trajectories of cognitive ability. Group-specific effects in univariate and bivariate model parameters were tested by comparing a model with all parameters free to vary across groups to models where the parameters were constrained to be group-invariant. The significance was then tested by comparing the log-likelihood difference (with an MLR correction for scaling factors) of each constrained model to the previous where the parameter was allowed to vary across groups. Proportional and coupling effects before and after 65 or 69 were tested together.

For processing speed, the linear decline was significantly steeper among those who developed dementia than those who remained cognitively intact (Table 2c right panel, dashed lines). No statistically significant group difference in rate of change was present for the other cognitive domains, though plots generally suggest a steeper decline in the dementia group (Figure 2a and b right panel, dashed lines). Thus, compared to the cognitively intact group, individuals wo were subsequently diagnosed with dementia had lower cognitive ability at later ages, although their estimated scores at age 50 were significantly higher for general cognitive ability and processing speed but lower for spatial ability and episodic (Table 2 and Table S1c, univariate estimates).

### 3.2 Bivariate trajectories of BMI and cognitive abilities

#### 3.2.1 Full sample

Comparisons of models with and without the coupling parameters are presented in Table 3. The relationships between BMI and general cognitive ability and BMI and processing speed were of a bidirectional nature, with cognitive ability driving change in BMI and BMI driving change in cognitive ability (Table 3). The relationship between BMI and spatial ability was of a unidirectional nature in the full sample, where spatial ability drives change in BMI, but BMI does not drive change in spatial ability. There was no statistically significant effect of coupling between BMI and episodic memory (Table 3), and no further tests were thus carried out to study the association. The model estimates for the full sample are presented in Table S1.

**Table 3:**
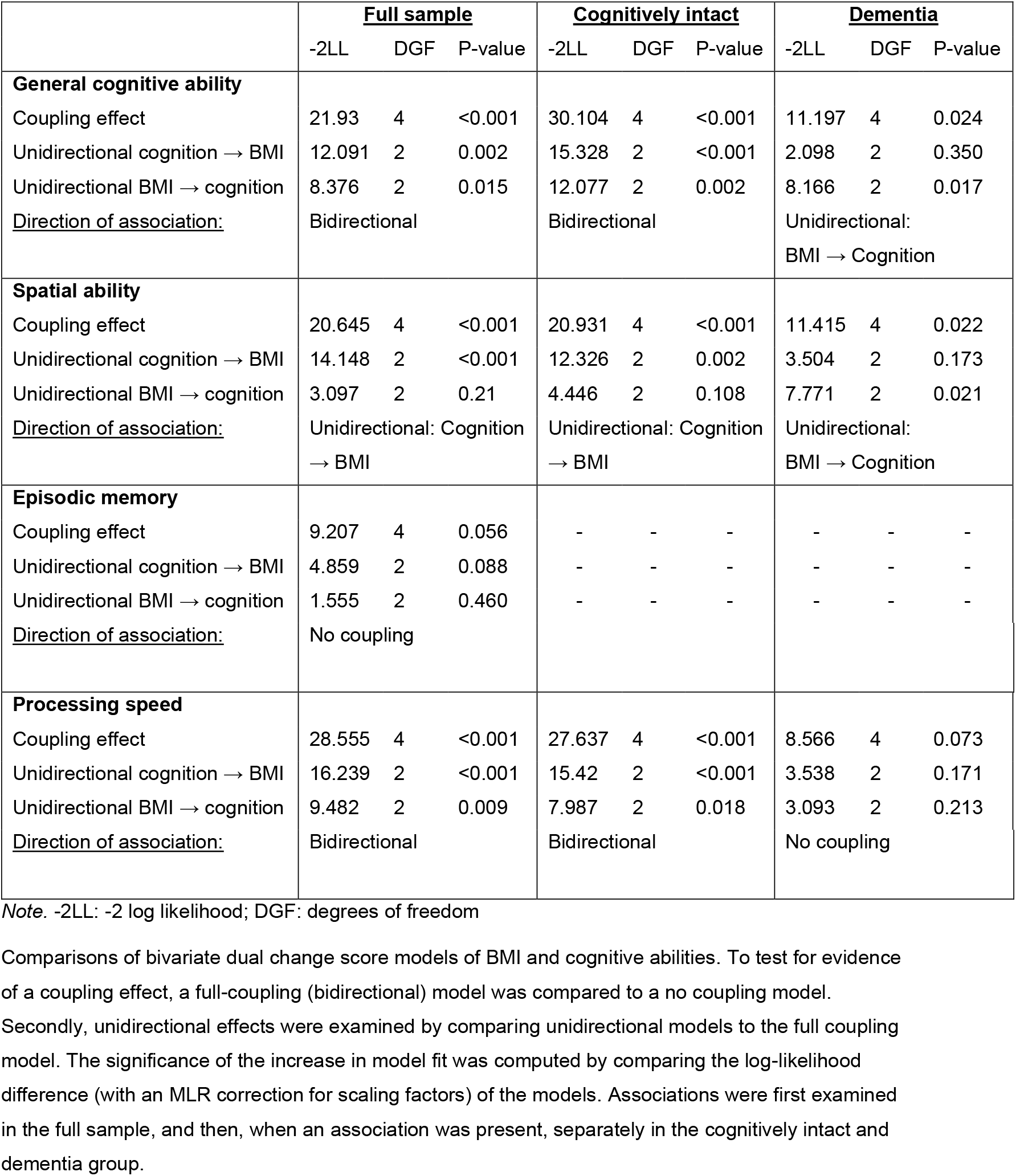
Bivariate model comparisons.

|  | <u>Full sample</u> |  |  | <u>Cognitively intact</u> |  |  | <u>Dementia</u> |  |  |
| --- | --- | --- | --- | --- | --- | --- | --- | --- | --- |
|  | -2LL | DGF | P-value | -2LL | DGF | P-value | -2LL | DGF | P-value |
| **General cognitive ability** |  |  |  |  |  |  |  |  |  |
| Coupling effect | 21.93 | 4 | <0.001 | 30.104 | 4 | <0.001 | 11.197 | 4 | 0.024 |
| Unidirectional cognition → BMI | 12.091 | 2 | 0.002 | 15.328 | 2 | <0.001 | 2.098 | 2 | 0.350 |
| Unidirectional BMI → cognition | 8.376 | 2 | 0.015 | 12.077 | 2 | 0.002 | 8.166 | 2 | 0.017 |
| <u>Direction of association:</u> | Bidirectional |  |  | Bidirectional |  |  | Unidirectional:<br>BMI → Cognition |  |  |
| **Spatial ability** |  |  |  |  |  |  |  |  |  |
| Coupling effect | 20.645 | 4 | <0.001 | 20.931 | 4 | <0.001 | 11.415 | 4 | 0.022 |
| Unidirectional cognition → BMI | 14.148 | 2 | <0.001 | 12.326 | 2 | 0.002 | 3.504 | 2 | 0.173 |
| Unidirectional BMI → cognition | 3.097 | 2 | 0.21 | 4.446 | 2 | 0.108 | 7.771 | 2 | 0.021 |
| <u>Direction of association:</u> | Unidirectional: Cognition<br>→ BMI |  |  | Unidirectional: Cognition<br>→ BMI |  |  | Unidirectional:<br>BMI → Cognition |  |  |
| **Episodic memory** |  |  |  |  |  |  |  |  |  |
| Coupling effect | 9.207 | 4 | 0.056 | - | - | - | - | - | - |
| Unidirectional cognition → BMI | 4.859 | 2 | 0.088 | - | - | - | - | - | - |
| Unidirectional BMI → cognition | 1.555 | 2 | 0.460 | - | - | - | - | - | - |
| <u>Direction of association:</u> | No coupling |  |  |  |  |  |  |  |  |
| **Processing speed** |  |  |  |  |  |  |  |  |  |
| Coupling effect | 28.555 | 4 | <0.001 | 27.637 | 4 | <0.001 | 8.566 | 4 | 0.073 |
| Unidirectional cognition → BMI | 16.239 | 2 | <0.001 | 15.42 | 2 | <0.001 | 3.538 | 2 | 0.171 |
| Unidirectional BMI → cognition | 9.482 | 2 | 0.009 | 7.987 | 2 | 0.018 | 3.093 | 2 | 0.213 |
| <u>Direction of association:</u> | Bidirectional |  |  | Bidirectional |  |  | No coupling |  |  |

Note. -2LL: -2 log likelihood; DGF: degrees of freedom
Comparisons of bivariate dual change score models of BMI and cognitive abilities. To test for evidence of a coupling effect, a full-coupling (bidirectional) model was compared to a no coupling model.
Secondly, unidirectional effects were examined by comparing unidirectional models to the full coupling model. The significance of the increase in model fit was computed by comparing the log-likelihood difference (with an MLR correction for scaling factors) of the models. Associations were first examined in the full sample, and then, when an association was present, separately in the cognitively intact and dementia group.

#### 3.1.1 By dementia status

Longitudinal trajectories of change in BMI and cognitive abilities, with and without considering the coupling parameters, are shown in Figure 2. The no-coupling trajectories correspond to the univariate trajectories, and the full-coupling trajectories show change in BMI when considering the effect of cognitive ability, and vice versa.

Trajectory estimates from bivariate models are presented in Table 2, with full model estimates presented in Table S1. Likelihood ratio tests (LRT) demonstrated significant group differences for the effect of general cognitive ability on change in BMI, and for the effect of BMI on general cognitive ability and spatial ability (Table 2).

##### Cognitively intact

Among individuals who remained cognitively intact, the nature of the relationships between BMI and cognitive abilities were the same as in the full sample, namely that the associations between BMI and general cognitive ability and BMI and processing speed were of a bidirectional nature, while spatial ability drives change in BMI but not the opposite (Table 3).

By comparing trajectory estimates from univariate and bivariate models, the effect of the coupling mechanisms can be studied. The coupling parameters should then be interpreted together with changes in the linear slope and proportional effects. When the effect of general cognitive ability, as well as that of spatial ability and processing speed, was considered, BMI at age 50 was lower, the negative linear slope steeper, and buffering from proportional effects after age 69 weaker (Table 2). However, there was an additional buffering effect from coupling parameters, both before and after age 69. As the coupling parameters are multiplied with level of the other variable at the preceding occasion, this means that higher cognitive ability predicts slight increases in BMI from age 50 to 69, and less decline in BMI after age 69. In Figure 2 (left panel), this is seen as steeper increase from age 50 to 69, followed by more stable level after age 69. However, Figure 2 shows a steeper decline at later ages when the effect of cognition is considered, likely due to lower levels of general cognitive ability limiting buffering from coupling effect.

When the effect of BMI was considered, general cognitive ability and processing speed were slightly higher at age 50, but the linear slope showed steeper decline (Table 2a and c). However, this was buffered by stronger proportional effects, especially before age 65. The coupling effects from BMI added to the decline before age 65, but buffered against it after age 65. In Figure 2 (right panel), this is seen as a slightly steeper decline in cognitive abilities from age 50 to 65, and slightly less decline from age 65 to 89 when the effect of BMI is considered.

The coupling effects of BMI on general cognitive ability and processing speed were negative from age 50 to 65, meaning that higher BMI predicts a steeper decline in cognitive ability, but positive from age 65 through 89, with higher BMI predicting less decline in cognitive ability (Table 2a and c). Taken together, this indicates that in older ages a stable BMI predicts a stable cognitive ability and vice versa.

##### Dementia

Among individuals who were later diagnosed with dementia the associations between BMI and general cognitive ability and BMI and spatial ability were unidirectional, with BMI driving change in cognitive ability, but not the opposite (Table 3). Both before and after age 69, higher BMI predicted steeper decline in cognition. This result is indicated by a positive overall linear slope in the bivariate model (in contrast to the univariate model, where the linear slope was negative; Table 2a and b), counteracted by strongly negative coupling effects (multiplied by BMI level at the preceding occasion) which thus drive the decline. The buffering from proportional effects were less affected by the influence of BMI. There was no statistically significant coupling present for the association between BMI and processing speed (Table 3).

As visualized in Figure 2, considering the effect of BMI led to a more stable cognitive ability from age 50 to 65, followed by comparable decline in general cognition but a steeper decline in spatial ability.

## 4. Discussion

We here used the powerful DCSM to study the dynamic relationship between BMI and cognitive abilities from age 50 through 89, and demonstrated that the nature of the association differed between individuals who remained cognitively intact and those who were later diagnosed with dementia. Among individuals who were subsequently diagnosed with dementia, higher BMI predicted steeper decline in general cognitive ability, and in spatial ability in particular, throughout midlife and late-life, while level of cognitive ability did not predict change in BMI. Among those who remained cognitively intact, for general cognitive ability, and processing speed in particular, the associations with BMI were of a bidirectional nature, such that higher BMI predicted a steeper decline in cognitive abilities before age 65 but—in contrast to the predementia group--buffered against decline after age 65. In turn, general cognitive ability, and particularly level of processing speed and spatial ability, drives change in BMI, with higher cognitive ability predicting more increase in midlife and less decline in late life. No coupling effect between BMI and episodic memory was identified.

The differences in the results by dementia status highlight that the relationship between BMI and cognitive abilities is dysfunctional in preclinical dementia. Among individuals who remain cognitively intact, stable cognitive abilities predict stability in BMI, and a stable BMI predicts stability in cognitive abilities in older ages. Among individuals who developed dementia, on the other hand, these stabilizing effects are missing. Instead, we see a decline in cognitive ability which is largely a function of the coupling effects from BMI, with too little buffering from cognitive level itself to compensate for the negative effects. This indicates that lower cognitive ability, per se, does not explain weight loss in preclinical dementia. Rabin and colleagues[19] demonstrated that a higher amyloid-beta burden predicted more decline in BMI in a sample of cognitively normal individuals at baseline. Importantly, the results persisted in models adjusted for cognitive performance. Müller et al. have shown that BMI also declines 10-20 years prior to the expected onset of autosomal dominant Alzheimer’s disease (AD)[20]. The decline began well before any clinical or cognitive symptoms were present, and is thus not an effect of cognitive impairment, but the authors did note that BMI was associated with lower cognitive performance, still within normal ranges. This also shows that decline in BMI is not only an effect of older age, but specifically linked to the preclinical AD process. Taken together, these findings and ours indicate that decline in BMI seen in the preclinical dementia phase is likely not driven by declining cognitive ability.

This study was built on a well-characterized sample with objectively measured BMI, robust cognitive measures, and dementia assessed during the study as well as through register linkage. The sample entailed us to study change from age 50 and through 89 and to apply the powerful DCSM. The study is not without limitations. The sample size, though phenotypically rich, was limited. As in all studies of older adults, poor health may lead to survival bias and attrition rate[23]. This issue may be more important among individuals later diagnosed with dementia, highlighting the importance of supplementing dementia diagnosed during the studies with register linkage after the end of follow-up. It should be noted, however, that while dementia information from healthcare registers has excellent specificity, the sensitivity is rather low[24], and some dementia diagnoses may have been missed.

In conclusion, we here show that the longitudinal association between BMI and cognitive abilities differ among individuals who develop dementia compared to those who remain cognitively intact. While BMI and cognitive abilities stabilize each other when cognition remains intact, this buffering effect is missing in the preclinical phase of dementia, where only a negative effect of higher BMI remains. This is in agreement with previous evidence, indicating that weight loss in preclinical dementia is not an effect of cognitive impairment, but may rather be an effect downstream of AD pathology[19, 20].

## Data Availability

The SATSA data are available in the at National Archive of Computerized Data on Aging under accession nunber ICPSR 3843 (https://www.icpsr.umich.edu/web/NACDA/studies/3843). The OCTO-Twin and GENDER data can be applied for through https://www.near-aging.se/. 
All codes used to generate analysis data and for conducting analyses are available upon request to the corresponding author.

## Acknowledgements

This work was supported by the Swedish Research Council for Health, Working Life and Welfare (2018-01201); the Swedish Research Council (2016-03081); and the National Institutes of Health (NIH AG060470).

We acknowledge the Swedish Twin Registry for access to data. The Swedish Twin Registry is managed by Karolinska Institutet and receives funding through the Swedish Research Council under the grant no. 2017-00641.

SATSA was supported by National Institutes of Health (NIH; grants AG04563 and AG10175), the MacArthur Foundation Research Network on Successful Aging, the Swedish Research Council for Working Life and Social Research (FAS; Grants 97:0147:1B, 2009-0795), and the Swedish Research Council (825-2007-7460 and 825-2009-6141). OCTO-Twin was supported by NIH (grant R01 AG08861). GENDER was supported by the MacArthur Foundation Research Network on Successful Aging, The Axel and Margaret Ax:son Johnson’s Foundation, The Swedish Council for Social Research, and the Swedish Foundation for Health Care Sciences and Allergy Research.

