## Supplementary material for "Change in cognition and body mass index in relation to preclinical dementia": Figure S1

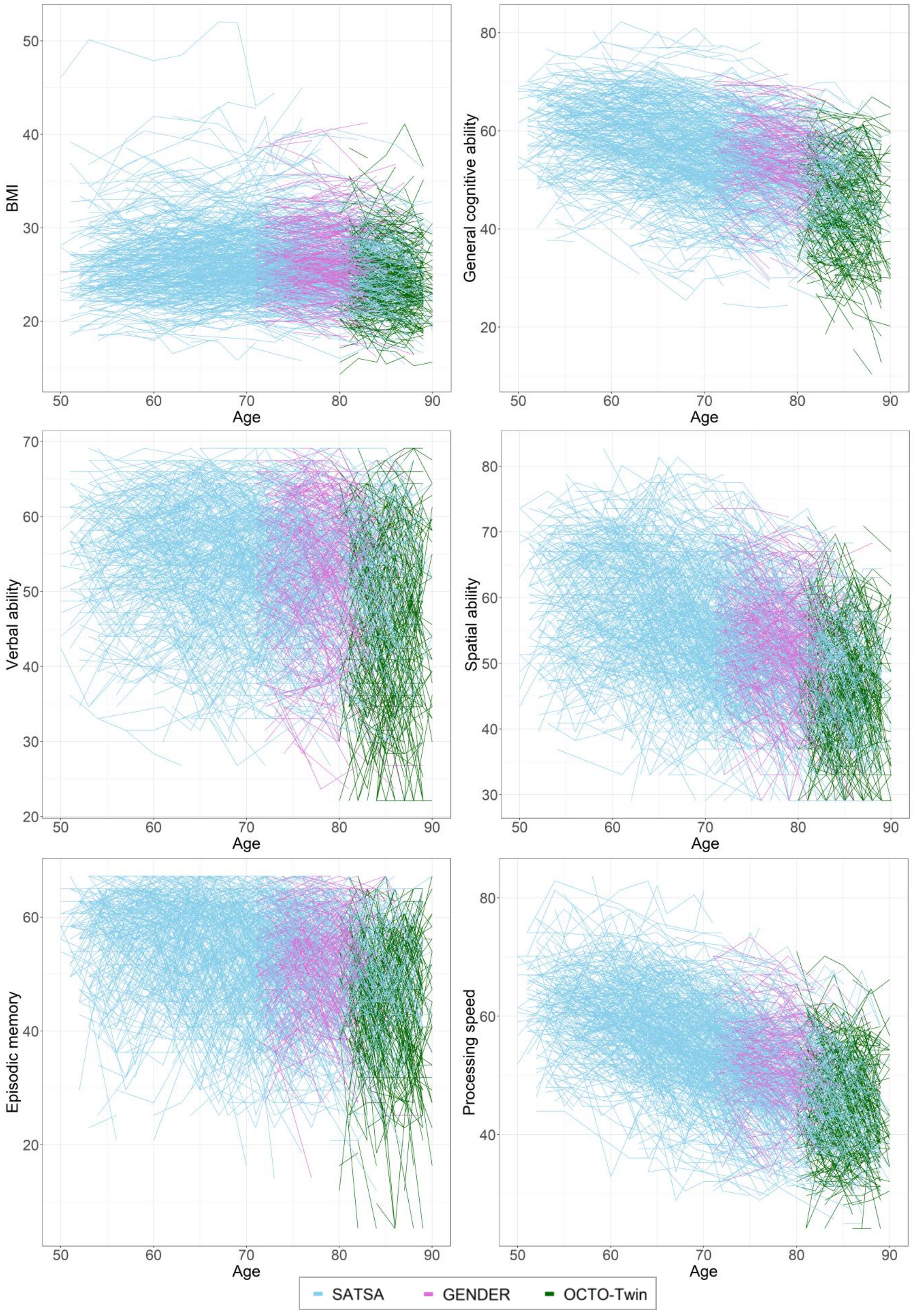


**Figure S1: Individual trajectories of body mass index and cognitive abilities from age 50-89, by study.** Individual values are plotted across age, with SATSA participant represented in blue, GENDER participants in purple, and OCTO-Twin participants in green.
